# Proteomics analysis in urinary bladder cancer patients identifies urinary SOD2 as a predictive marker of recurrence

**DOI:** 10.1101/2021.12.13.21267125

**Authors:** Nitu Kumari, Subasa Chandra Bishwal, Shweta Chaudhary, Deepankar Malalkar, Uma S. Dubey, Pawan Vasudeva, Anup Kumar, Sunita Saxena, Ranjan Kumar Nanda, Usha Agrawal

## Abstract

Early non-invasive detection of tumor is an urgent clinical need for managing urothelial bladder cancer. Cystoscopy and cytology are the current standards for diagnosis of recurrence, but are limited by low sensitivity. Quantitative proteomics tool was employed to identify important deregulated molecules in bladder cancer tissues and validated using Western blot and immunohistochemistry analysis. A set of 1137 proteins were identified in four paired bladder cancer patients. Among these, 64 proteins were deregulated in all cases among which 9 were commonly up-regulated. The Ingenuity Pathway Analysis (IPA) generated top 11 Networks in which three commonly upregulated (SERPING1, SOD2 and HSPB6) proteins were involved and selected for further validation. Tissue expression of SOD2, SERPING1 and HSPB6 monitored in an independent sample set (n=18) by immuno-histochemical analysis showed similar profile. Western blot analysis of these proteins in urine of bladder cancer (n=26) and healthy subjects (n=10) showed a specificity and sensitivity of >80% for SOD2 and so was selected for further validation in a separate set (n=150) by ELISA. Significant elevation in urinary SOD2 level was found in urothelial bladder cancer patients compared to healthy controls and in recurrent cases compared to primary (p-value<0.001). Kaplan Meier survival analysis showed urinary SOD2 concentration >2,100 pg/ml was significantly associated with poorer survival.Cumulative survival of patient with low SOD2 concentration was 34.4% compared to 18.9% in patient with high SOD2 at 24 months (p=0.025). The study identifies SOD2 as a non-invasive biomarker which may help to extend the period between cystoscopies during follow-up.

**Significance:** Cystoscopy is an invasive and painful method commonly used for diagnosis of urothelial bladder cancer. Non-invasive methods having high specificity and sensitivity to monitor the patients for recurrence are unavailable. Our study reveals significantly higher SOD2 level in drug naive and reoccurring bladder cancer tissues, and similar profile was observed in the parallel urine samples. Hence, SOD2 seems to be a useful biomarker of recurrent urothelial bladder cancer and predict the survival of patients.

## Introduction

Urinary bladder cancer affects millions of people annually with ∼573,278 new cases reported in 2020.^1^ It is the tenth most common malignancy with the highest rate of recurrence amongst all cancer. In US and European union alone, the estimated costs associated with bladder cancer management were $4 billion and €4.9 billion respectively.^2,3^ Majority of these newly diagnosed cases present with macroscopic haematuria. Initial investigation involves cytoscopy, urine cytology and cases are confirmed after transurethral resection of bladder tumour (TURBT). Cystoscopy is an invasive procedure, whereas urine cytology is limited by the low sensitivity (4–31%).^4^ So, an early non-invasive screening method to predict recurrences will improve bladder cancer disease management and associated costs.

Out of several risk factors, tobacco smoking accounts for 50% of total bladder cancer cases followed by environmental and occupational exposures to aromatic amines, polycyclic aromatic and chlorinated hydrocarbons.^5,6^ Additionally, sex-steroid axis and gender-specific risk factors play critical role in bladder cancer development and progression. In male, the case incidence is threefold higher whereas more advanced stage tumours are reported in females.^7^ Early diagnosis is the key for efficient management of this critical condition. FDA approved urine-based surveillance test as adjunct to cytoscopy aid early diagnosis but present with modest sensitivity (55-70%) and specificity (71-83%).^8,9^ Furthermore, false positive results due to commonly observed haematuria and inflammation conditions other than urinary bladder cancer may limit its utility.

In this study, we profiled the perturbed proteome of adjacent bladder cancer and healthy tissues to identify important deregulated tissue specific putative markers. Further, urine levels of these markers were monitored to calculate their predictive accuracy to predict disease recurrence.

## Materials and Methods

### Subject recruitment for sample collection and classification

All study subjects provided signed informed consent to participate in this study following the approval of the Ethics Committee of V.M.Medical College & Safdarjang Hospital New Delhi (No. 52-11-EC(5/17) dated 1st March 2012). Demographic details of the study subjects are summarized in Table S1. Paired tissue biopsy samples from cancer and nearby normal mucosa were collected by transurethral resection of bladder tumour (TURBT) and radical cystectomy from the patients. After excision, a part of tissue piece was stored in 10% formalin and embedded in paraffin (FFPE tissue) for immuno-histopathological (IHC) analysis and rest stored at -80°C until proteomics analysis. The cases were classified by histo-pathological examination into low- and high-grade cancer cases (L/H) and stages (S1/2) following the World Health Organization/International Society of Urological Pathology (WHO/ISUP) consensus classification of urothelial neoplasms. Based on the case history, the subjects were grouped as recurrent (R) or non-recurrent (NR) groups. First morning mid-stream urine specimens were collected prior to surgery and from age-gender matched control subjects to use in validation phases. Protease inhibitor mixture (P8340, Sigma, USA) and sodium azide (1 mM) were added to urine. The samples were centrifuged at 5,000 × *g* for 30 min at 4°C within 3 hours of collection. The supernatants were stored at -80°C in aliquots for Western blot and ELISA analyses. Independent sample sets were used in discovery (tissue sample, n= 8 using mass spectrometry/iTRAQ LC-MS/MS) and validation phases (FFPE tissue sample, n=18 using IHC; urine sample, n=36 using Western blot and urine sample, n=150 using ELISA).

### Tissue sample processing for quantitative proteomics experiment

Stored tissue samples were homogenized using liquid nitrogen and mixed in minimum volume of RIPA buffer (Thermo Fisher Scientific, USA) supplemented with protease inhibitor (P8340, Sigma, USA). The homogenates were sonicated (80Hz, 2 cycles for 30 sec in ice) and supernatant was collected at 13,000 × *g* for 20 min at 4°C. Concentration of total protein was determined by a BCA protein assay (23225, Thermo Fisher Scientific, USA) following the manufacturer’s instruction. Isolated tissue proteins from all samples (low grade technical replicate-1: LGT1, high grade technical replicate-1 responder: HGT1R, high grade technical replicate-1 non-responder: HGT1NR and high grade technical-2:HGT2) to be used for proteomics experiments were probed by running in 12% SDS-PAGE and the gel was silver stained (Figure S1A). Tryptic peptides were generated from equal amount of protein (∼100 µg) from each study groups (LGT1, HGT1R, HGT1NR and HGT2) and labelled with isobaric tags following manufacturer’s instructions (iTRAQ 8-plex, AB Sciex, USA). Briefly, protein samples were precipitated with pre-chilled acetone (v/v: 1/6) and incubated at -80°C overnight. Precipitated proteins were collected after centrifugation at 10,000 × *g* for 30 min at 4°C and pellet was reconstituted in dissolution buffer (20 μl, triethylammonium bicarbonate, 0.5 M, pH 8.5) and denaturant buffer (SDS, 0.05 % w/v). Tissue proteins after reduction and cysteine block were digested with trypsin (10 μl, from iTRAQ 8plex kit) and incubated at 37°C overnight. Resulting tryptic peptides were dried in a SpeedVac (Labconco, USA) and reconstituted in dissolution buffer (pH ∼8.0). Tryptic peptides from all samples were labelled with isobaric tags and after incubation pooled together to dry using a SpeedVac. Samples from tumour (pTa stage: non-invasive) and adjacent normal tissues were labelled using 113–116 tags, whereas for the invasive tumours (pT2: invasive) 117–119 and 121 tags were used (Figure 1, Figure S1).

**Figure 1.**
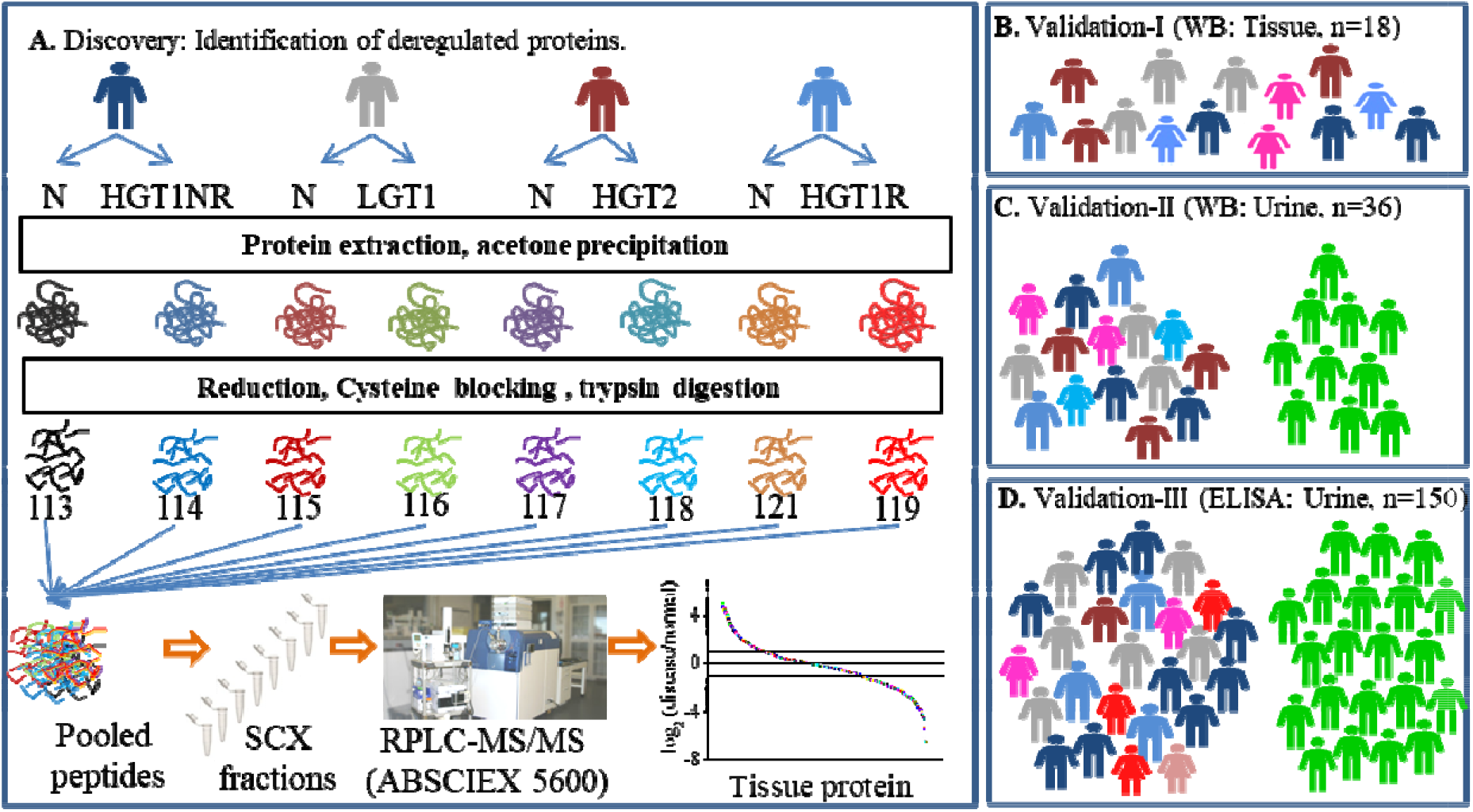
Schematic workflow used to identify deregulated molecules in cancer tissues of bladder cancer patients. A) Paired samples (n=4, all male) of bladder cancer patients including High grade stage 1 non recurrent (HGT1NR), low grade stage 1 (LGT1), High grade stage 2 (HGT2), High grade stage 1 recurrent (HGT1R) tumor tissue and adjacent mucosa (N) confirmed by histopathology were used for protein extraction and labelling of tryptic peptides for mass spcetrometry analysis. B) and C). The selected candidates were evaluated in both tissue and urine samples of bladder cancer patients by immunohistochemistry (localization of protein) and Western blot analysis in two verification phases. D) Validation of one marker molecule was performed in urine of an independent sample set (n=150) by ELISA.

### Tryptic peptide fractionation and mass spectrometry data acquisition

The pooled labelled tryptic peptides were dissolved in ammonium formate buffer (5mM) in acetonitrile (ACN, 30%) with formic acid (FA, 0.1%) and fractionated using ICAT™ (ABI, USA) cartridge. Briefly, the cartridge was conditioned with ammonium formate (500 mM) in ACN (30%) with FA (0.1%) and peptide fractions were eluted using increasing buffer concentration (30 mM, 80 mM, 120 mM, 180 mM, 250 mM, 300 mM, 400 mM and 500 mM) with a flow rate of single drop/sec. This resulted in a total of 8 fractions and each fraction was dried using a SpeedVac. The solvent (mixture of 2% Acetonitrile and 0.01% Formic acid) was used to dissolve each peptide fraction (∼32.5 μg) and 6 µl peptides (approximately 1.95µg) were loaded onto a reverse phase peptide Eksigent ChromoXP trap (200 mm × 0.5 mm, 3 mm, 120 Å) column. Peptides were desalted at a flow rate of 3 ml/min for 45 mins. Desalted peptide was seprated at a flow rate of 225 nl/min using 90 minutes of linear gradient of 5% to 90% solvent B (98% ACN with 0.1% FA) using an Eksigent C18 column (75 mm × 15 cm, 3 mm, 100 Å). Samples were analyzed using a TripleTOF5600 (Sciex, USA) MS coupled to an Eksigent NanoLC-Ultra 2D plus system. Analysis of sample uses information dependent acquisition mode with a TOF MS survey Scan (3501250 m/z), which uses maximum of 12 precursor ion/cycle with an accumulation time of 250 ms were selected for peptide fragmentation. Setting was used as nebulizing gas (25), curtain gas (25), a heater interface temperature (130°C) and an ion spray voltage 2600V for analysis of sample. Each MS/MS spectrum was accumulated at 1.5 sec of cycle time for 100 ms (100-1500 m/z) and acquired using iTRAQ reagent settings (high sensitivity mode for adjust collision energy). The threshold precursor ion intensity >120 cps and charge state from +2 to +5 and exclusion period 10 sec were selected for MS/MS fragmention of only the parent ions.

### Mass Spectrometry Data Availability and Important Protein Identification

The Raw MS spectra data were processed using Analyst (Sciex) to extract mass spec files (.wiff) and all raw data files have been deposited via the PRIDE partner repository to the ProteomeXchange Consortium with the dataset identifier PXD007070.^23^ These wiff files were processed using ProteinPilot (version 5.0, Sciex) software to identify peptides and proteins at a false discovery rate (FDR) of 2% at peptide and protein labels. Proteins were identified on the basis of having unused score of >1.3 and at least 2 or more identified peptides. The bias correction and background correction options were executed. Protein relative expression ratios were based on the peak area ratios of the peptides from the same protein and the resulting dataset was auto bias-corrected to eliminate any variability. Fold changes are presented in log_2_(cancer/normal) and a fold change of >±1 were selected as important molecules. Gene ontology (GO) analysis was performed using Protein Analysis THrough Evolutionary Relationships (PANTHER) database v 6.1 (www.pantherdb.org). Pathway and network analysis were performed using Ingenuity Pathway Analysis of Differentially regulated proteins (Ingenuity Systems, Inc., Redwood City, CA).

### Immunohistochemistry analysis

Paraffin sections of cancerous and adjacent normal mucosa tissue samples from patient groups (low grade stage 1:LS1, high grade stage 1:HS1, high grade stage 2:HS2; 5 each; n=15), not used in proteomics experiment, were subjected to immunohistochemistry and the expression and localization of markers (SERPING1, HSPB6and SOD2) were probed. Briefly, FFPE tissues were dewaxed (in Xylene) and rehydrated in alcohol gradient (100%, 90%, 70%, 50% and water). Antigen retrieval was carried out using tris-EDTA buffer (pH 9.8) for 20 min at 95°C in a water bath. The samples were treated with hydrogen peroxide (H_2_O_2_; 3%) in methanol for 10 min and blocked using a blocking buffer (5% BSA in TBS) for 30 mins followed by overnight incubation with primary antibody [primary antibody Rabbit antiSERPING1 (Cat# PA5-13627 ThermoScientific, 1:100), Mouse antiSOD2 (Cat# sc-133134,Santa Cruz, 1:200), Mouse antiHSP20 (Cat# sc-133677,Santa Cruz, 1:100)].The same primary antibody was used in Western blot experiment. After washing with TBST, labelled secondary antibody (Super Sensitive™ Polymer HRP Kit, catalog# QD420-YIKE, BioGenex) was added and incubated for at least 2 hours. Tissue sections were stained for 2-3 minutes with 3,3’-diaminobenzidinetetrahydrochloride (DAB; 0.05%) containing H_2_O_2_ (0.01%). Sections were counterstained with Mayers’ haematoxylin solution, dehydrated and mounted. Slides were examined using an optical digital microscope (Zeiss, Germany) by independent authors. Based on the presence or absence of blot signal, expression of these proteins was scored as positive or negative respectively.

### Western blot analysis

Important deregulated proteins identified from the proteomics experiments were validated using a separate set sample from patients not used in the discovery phase by Western blot analysis. Briefly, urine protein samples (20-25 µg) were denatured in Laemmli-SDS-sample buffer and subjected to polyacrlyamide gel electrophoresis (12%) and subsequently transferred to nitrocellulose membranes (Millipore). After blocking the transferred blots with bovine serum albumin (5% BSA) for overnight, blots were incubated with primary antibody Rabbit antiSERPING1 (ThermoScientific, Cat# A18903, 1:1000), Mouse antiSOD2 (Santa Cruz, Cat # A-10648, 1:10000) and Mouse antiHSP20 (Santa Cruz, Cat # A-10648, 1:10000) for 2-3 hours at RT on a shaker. Membranes were incubated with the HRP-coupled secondary antibodies (Biogenex, Cat # SKU: QD400-GP) for 2 hours at room temperature on shaker. Tris buffer saline with Tween 20 (0.05 %) was used as washing buffer between each incubation step for 5 mins and the membrane was visualized using Chemiluminescent ECL agent (Cat# WBKLS0500 Millipore).

### ELISA to estimate urine SOD2 levels

Stored urine samples of the study subjects, were thawed on ice and after bringing to room temperature, SOD2 level was quantified using ELISA kit (R&D Systems, Cat#DYC3419). Briefly, 100 µl urine sample was taken in duplicate onto the coated plate and incubated for 2 hours at room tempeture. Aspirate each well and wash with Wash Buffer followed by addition of secondary antibody (20 min room temperature), substrate (20 min room temperature) and Stop solution following the manufacturers’ instructions. Optical density of each well were measured immediately, using a microplate reader set to 450 nm for SOD2 quantification.The absolute levels of urinary SOD2 protein were determined using standard curves run on each ELISA plate, and normalized by urine creatinine concentration (Creatinine Assay Kit, Abcam, Cat# ab65340).

### Statistical Analysis

The Mann Whitney test was used to evaluate the significance of differences in marker concentration between study groups. Median concentration of urinary marker was taken as a cut-off for survival analysis. Kaplan Meier analysis for recurrence-free survival was done and significance was computed by log-rank. A p-value of <0.05 was considered statistically significant. The social sciences (SPSS) software version 19 (SPSS, Chicago, IL, USA) was used for all statistical analyses.

## Results

### Bladder Cancer Tissue Proteome Show Stage Specific Deregulation

Comparative relative proteomics experiment including tissue proteins isolated from paired cancer and normal mucosa, identified a total of 1455, 1398, 1137 and 1399 proteins in LGT1, HGT1R, HGT1NR and HGT2 tumour tissues respectively (Figure 2A and 2B). Majority of the identified proteins showed similar abundance between conditions with a minor sub-set of significantly deregulated proteins. The recurrent tumour presented with 74 unique proteins (Figure 2B). From the common set of deregulated proteins (n=64) observed in all study groups, 9 were up-regulated and 19 down-regulated in urothelial cancerous tissues with respect to the healthy controls (Figure 2C, Table S2).

**Figure 2:**
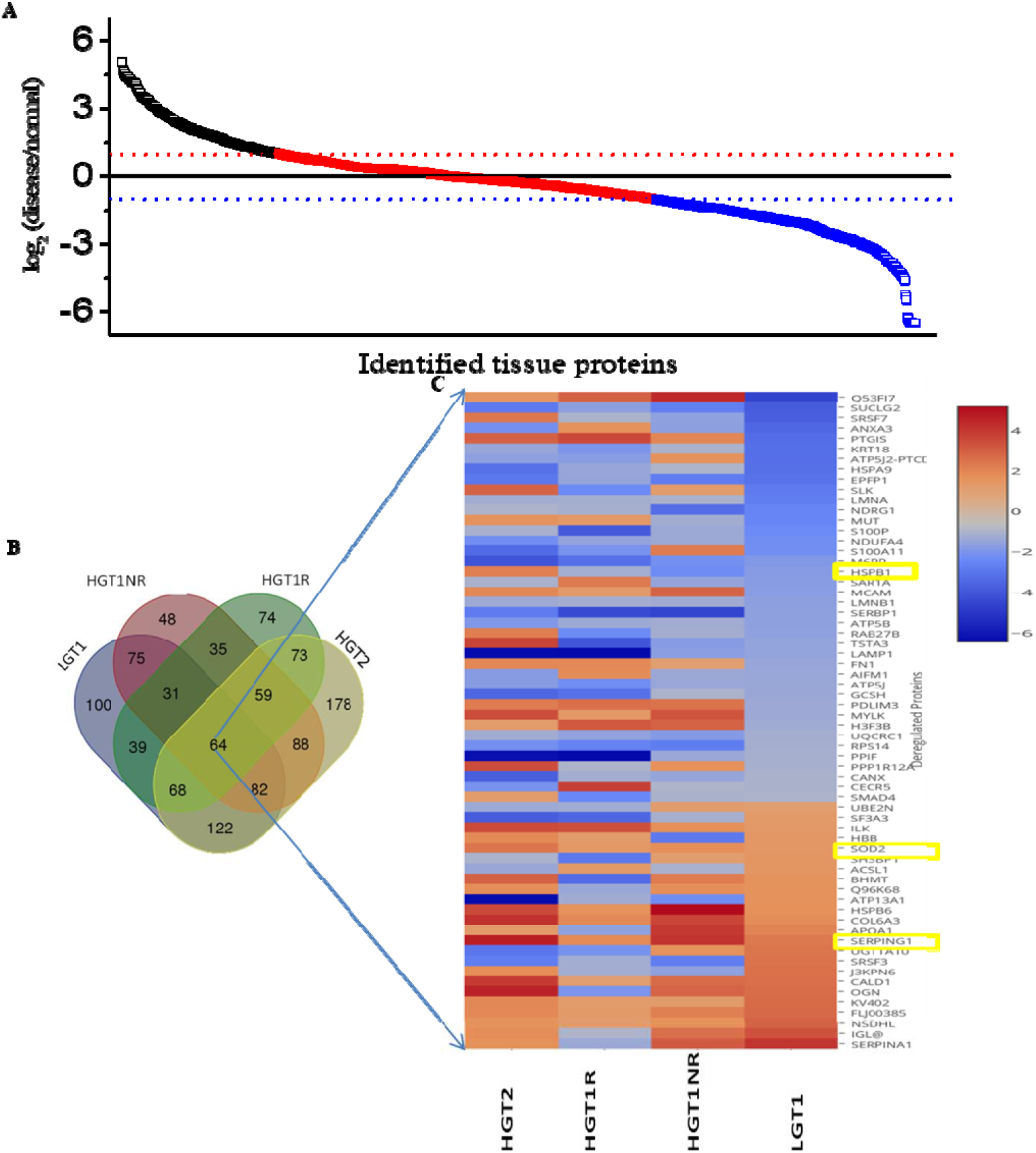
Selection of a set of important deregulated proteins from the discovery phase. A) A representative scatter plot depicting the identified proteins and their fold change in bladder cancer condition with respect to paired healthy tissue. B) Venn diagram presenting unique and common to sub groups of urothelial bladder cancer patients. C) Heat map showing deregulated profile of 64 common proteins in subgroups (LGT1, HGT1NR, HG1R and HGT2) of bladder cancer. Proteins are presented in row and subgroups of bladder cancer presented in colons. Color gradient show fold change and blue color represents minimum and red the maximum change.

### Pathway Analysis by DAVID

Gene Ontology (GO) terms determine the structure, localization and function. Within the Cellular component the differential proteins majorly grouped to 8 categories (Figure S2A). A set of 13 biological (Figure S2B) and 9 sets of molecular function enrichment was observed (Figure S2C).The Ingenuity Pathway Analysis (IPA) Core Analysis generated the top 11 Networks and revealed top three interactions between 24, 14 and 11 deregulated proteins. These proteins seem to be involved in cellular movement, haematological system, immune cell trafficking, nucleic acid metabolism, small molecule biochemistry, development disorder, cell death and survival, cellular development, cellular growth and proliferation (Table S3). Out of the up-regulated proteins, a signature consisting of SERPING1, HSPB6 and SOD2 were, part of the top three networks, present in all cancer tissues and identified as putative marker candidates for further validation.

### Diagnostic performance of the putative markers in tissue specimen

Immuno-histochemistry analysis of FFPE tissue samples showed similar cytoplasmic and membranous expression of SERPING1 in different grades and stages of tumour cells in all subgroups of bladder cancer patients (Figure 3A). SERPING1 was also found positive in lymphocytes (Figure 3A). HSPB6, a stress protein showed cytoplasmic and membranous expression selectively in tumour cells and was similar in studied grades and stages (Figure 3A). Cytoplasmic and membranous expression of SOD2 was seen in all grades and stages of bladder cancer. SOD2 expression was observed in the invasive front of tumour in HS2 but limited difference in localization was observed in low grade tumour (Figure 3A).

**Figure 3:**
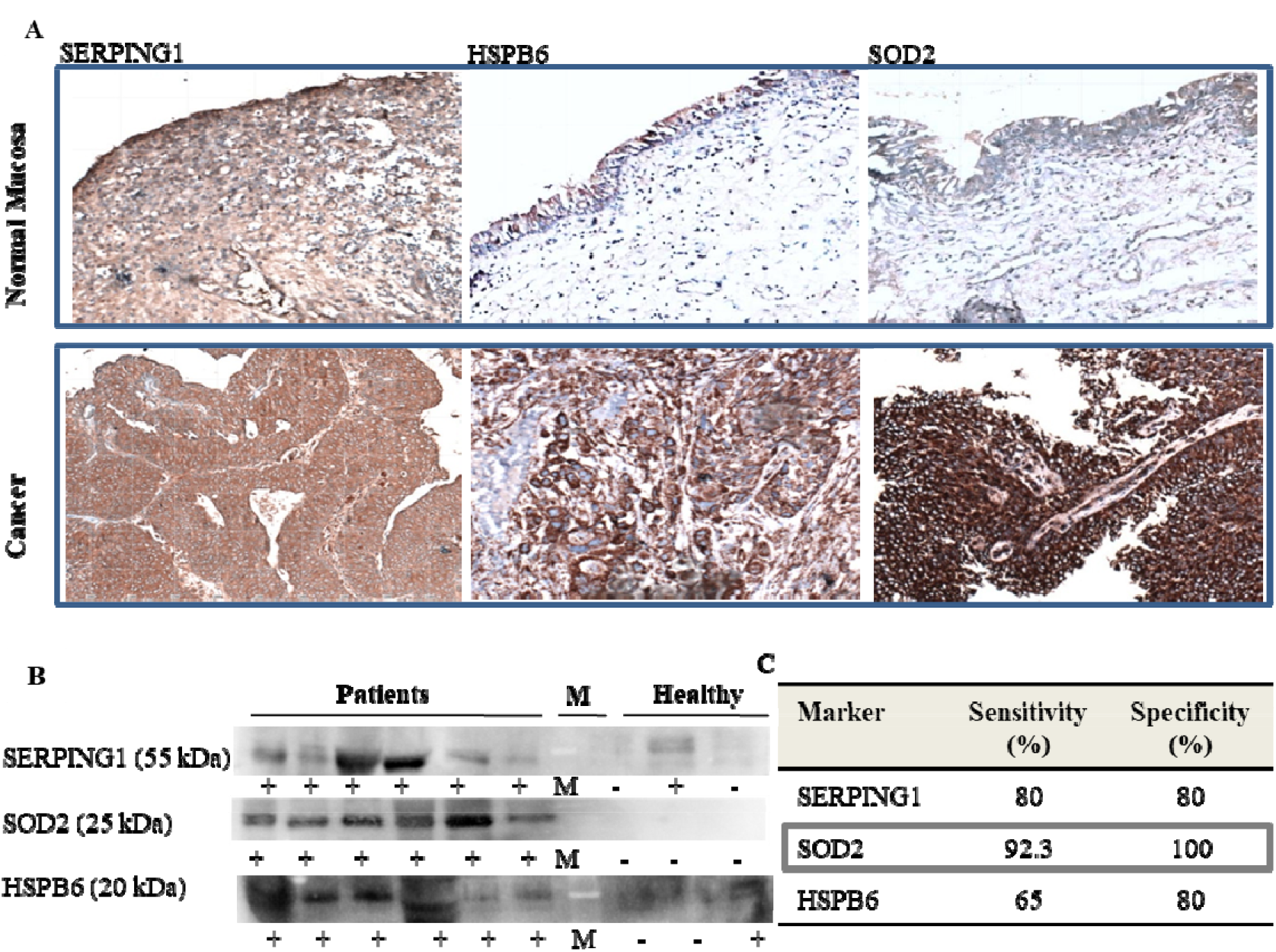
Verification of biomarkers in tissue and urine by immunohistochemistry and Western blot analysis respectively. A) Representative immunohistochemistry images of each marker proteins in adjacent normal mucosa in panel 1 and tumor tissue in panel 2 are shown. B) Representtaive Western blot images of each marker protein in urine samples from patients, marker (M) and healthy subjects is presented. Presence of the marker is pesented with ‘+’ and negative as ‘-’. C). Sensitivity and specficity calculated from the Western blot analysis of urine samples collected from 26 cancer patients and 10 controls. SERPING1 showed positive expression in cytoplasm of A) Normal mucosa, low grade non-muscle invasive urothelial bladder cancer captured at 20×, Urinary HSPB6 showed expression of HSPB6negative in adjacent mucosa, cytoplasm positivity in high grade non-muscle invasive urothelial bladder cancer captured at 20×; Immunohistochemistry for SOD2 showed negative expression in adjacent normal mucosa, cytoplasm of tumor cells are positive in low grade non-muscle invasive urothelial bladder cancer.

### Diagnostic Performances of the putative markers in Urine

The putative markers were further monitored in a set of urine samples (Validation-II; n=36; 26/10: urothelial cancer/healthy) by Western blot analysis. SERPING1 protein was detected in urine samples of majority of the bladder cancer patients (21 out of 26) and in healthy subjects (2 out of 10). The calculated sensitivity and specificity of urine SERPING1 was 80% (Figure 3B and 3C). Urine HSPB6 was identified in majority of bladder cancer patients (17 out of 26) and in a small group of healthy subjects (2 out of 10), resulting a sensitivity and specificity of 65% and 80% respectively (Figure 3B and 3C). SOD2 was identified in urine of all most all of bladder cancer patients (24 out of 26 urine), none in healthy with a calculated sensitivity of 92.3% and specificity of 100 % (Figure 3B and 3C).

Based on this data, absolute urine SOD2 levels was quantified in a separate set of subjects (Validation-III; n=150; bladder cancer/healthy: 100 (41 recurrent/50)). Significantly high urinary SOD2 level was observed in bladder cancer patients compared to healthy controls (p<0.001) (Figure 4A). Further sub-grouping of these test subjects as primary and recurrent groups showed elevated median concentration of urinary SOD2 in recurrent groups (Figure 4B). Median urinary SOD2 (2,100 pg/ml) was taken as cut off value and Kaplan Meier survival analysis showed that above this urine SOD2 levels were significantly associated with poorer recurrence-free survival. The cumulative survival of patient with low SOD2 concentration was 34.4% compared to 18.9% in patients having high urinary SOD2 at 24 months (p<0.025) (Figure 4C).

**Figure 4:**
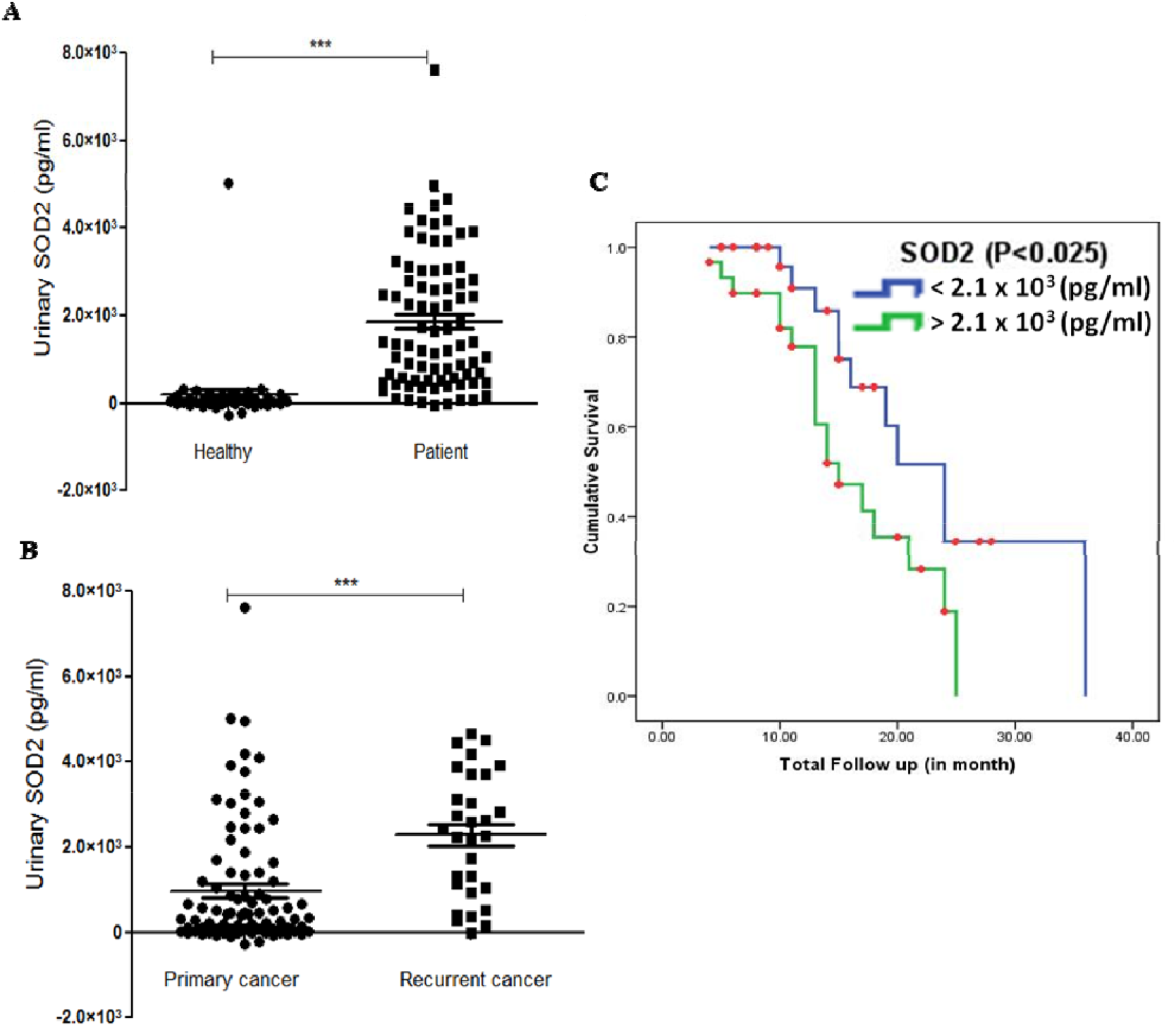
Higher urinary SOD2 level was obsered in fresh and recurrent bladder cancer patients. A) Significantly higher urinary SOD2 concentration was observed in primary bladder cancer patients with respect to healthy controls (p-value < 0.001, Mann Whitney U). B) In recurrent bladder cancer patients higher levels of urinary SOD2 levls with repsect to primary cases (p-value < 0.001) was observed. C) Kaplan Meier analysis showed higher urinary SOD2 (>2100pg/ml) was significantly associated with poorer survival of bladder cancer patients (log-rank t-test, p<0.025).

## Discussion

Early diagnosis of new and recurrent bladder cancer cases is of key importance and monitoring putative markers preferably in non-invasive diagnostic matrix like urine might significantly improve patient outcome. Dysregulated molecular patterns have been reported in the cancerous tissues of various organs like lungs, pancreas and colon.^10,11,12^ Elucidating genomic variations in cancer specific tissues proved useful for early diagnosis and personalized therapy development.^13^

In this study, using paired urothelial tumour and adjacent normal tissues and a multiplex iTRAQ experiment, a set of 28 deregulated molecules (9 up-regulated) in all grades and stages of bladder cancer was identified. IPA analysis revealed a complex network with SERPING1, HSPB6 and SOD2 involved in the topmost scored network. Higher abundance of these putative marker molecules was observed in the cancerous tissues. Immuno-histochemistry analysis of a separate validation set samples confirmed the altered expression of putative markers observed from the proteomics experiment. Interestingly, a recent report showed that healthy tissues isolated from cancer patients showed perturbed intermediate molecular profile from healthy controls.^14^ Comparative molecular profiling of cancerous and healthy tissues harvested from different case and control subjects were commonly followed. However, it will include variations contributed from environment and other host genetic factors and the adopted parallel tissue sampling procedure provide disease relevant variations. The putative tissue specific markers SERPING1, HSPB6 and SOD2 were monitored in urine samples of an additional validation set and SOD2 levels showed highest sensitivity and specificity values. Additionally, urine SOD2 levels were significantly higher in the recurrent bladder cancer patients from the time of primary diagnosis. Higher urinary SOD2 levels (>2,100 pg/ml) was found to be associated significantly with the disease recurrence and poor recurrence-free survival.

SOD2 is a member of iron/manganese superoxide dismutase family and binds to superoxide by-products released from mitochondrial electron transport chain or oxidative phosphorylation to transform them to hydrogen peroxide and diatomic oxygen.^15^ It clears mitochondrial reactive oxygen species (ROS) and protect against cell death. It has an anti-apoptotic role against oxidative stress, inflammatory cytokines and ionizing radiation. SOD2 exhibits anti-tumour as well as pro-tumour roles depending upon the type and stages of tumour.^16-19^ Reduced SOD2 expression results in DNA damage with increased cell proliferation leading to tumour genesis whereas increased SOD2 expression during later stage of tumour has been shown to assist metastasis.^17^ Higher SOD2 levels were also reported in penile, cervical, gastric/esophageal, colorectal and lung cancer cells.^20-24^ Higher SOD2 expression in stage III cervical carcinoma showed strong correlation with tumour recurrence.^25^ Over-expression of SOD2 was also reported in metastatic bladder cancer cell line compared to non-metastatic ones.^26,27^ Earlier reports showed association between SOD2 polymorphism and bladder cancer risk in different populations and needs further validation.^28,29^

However, limited reports on urine SOD2 level as a marker of urinary bladder cancer patients, as reported in this study, are available. The sensitivity calculated from the SOD2 expression levels in bladder cancer tissues was >90% and higher sensitivity (93% and 100% specificity) reflected from urine analysis. To the best of our knowledge, high urinary SOD2 levels to predict recurrence and poor recurrence-free survival in urinary bladder cancer patients reported in present study is first of its kind. Furthermore, urinary SOD2 concentration could be measured in patients having other genitourinary problems or benign conditions of the urinary tract such as renal stones, infection, and hematuria to evaluate appropriate predictive ability of this identified marker. It will be interesting to validate these observations in bladder cancer patients from other clinical settings preferably from the developed world. After further validation, SOD2 may find its usefulness as a surveillance marker for triaging urothelial bladder cancer patients for subsequent invasive cystoscopy tests for confirmation.

Biomarkers that represent highly sensitive and specific indicators of disease could be useful to measure treatment effectiveness, and non-invasive marker-based tests provide patient convenience. Few limitations in this study includes limited sample size which needs additional validation in larger cohort to carefully predict the potential of SOD2 as a prognostic factor in urinary bladder cancer. In addition, the present study has concentrated on a few of the markers which are up-regulated and the markers which are down-regulated were not validated owing to their lower expression level which may be below the lower limit of detectionin urine. Hence future studies on all the differentially regulated proteins may yield a better panel of biosignature with higher sensitivity and may elucidate the patho-physiological changes at pathway levels for designing targeted therapies.

## Data Availability

All data produced are available online at ProteomeXchange Consortium with the dataset identifier PXD007070.

## Acknowledgements

The study was supported by core funding from National Iinstitute of Pathology to UA and from International Centre for Genetic Engineering and Biotechnology New Delhi to RKN. NK was supported by fellowship from Indian Council of Medical Research, Government of India. SC is a Shyama Prasad Mukherjee fellow supported by Council of Scientific and Industrial Research, Government of India.

## Graphical Abstract

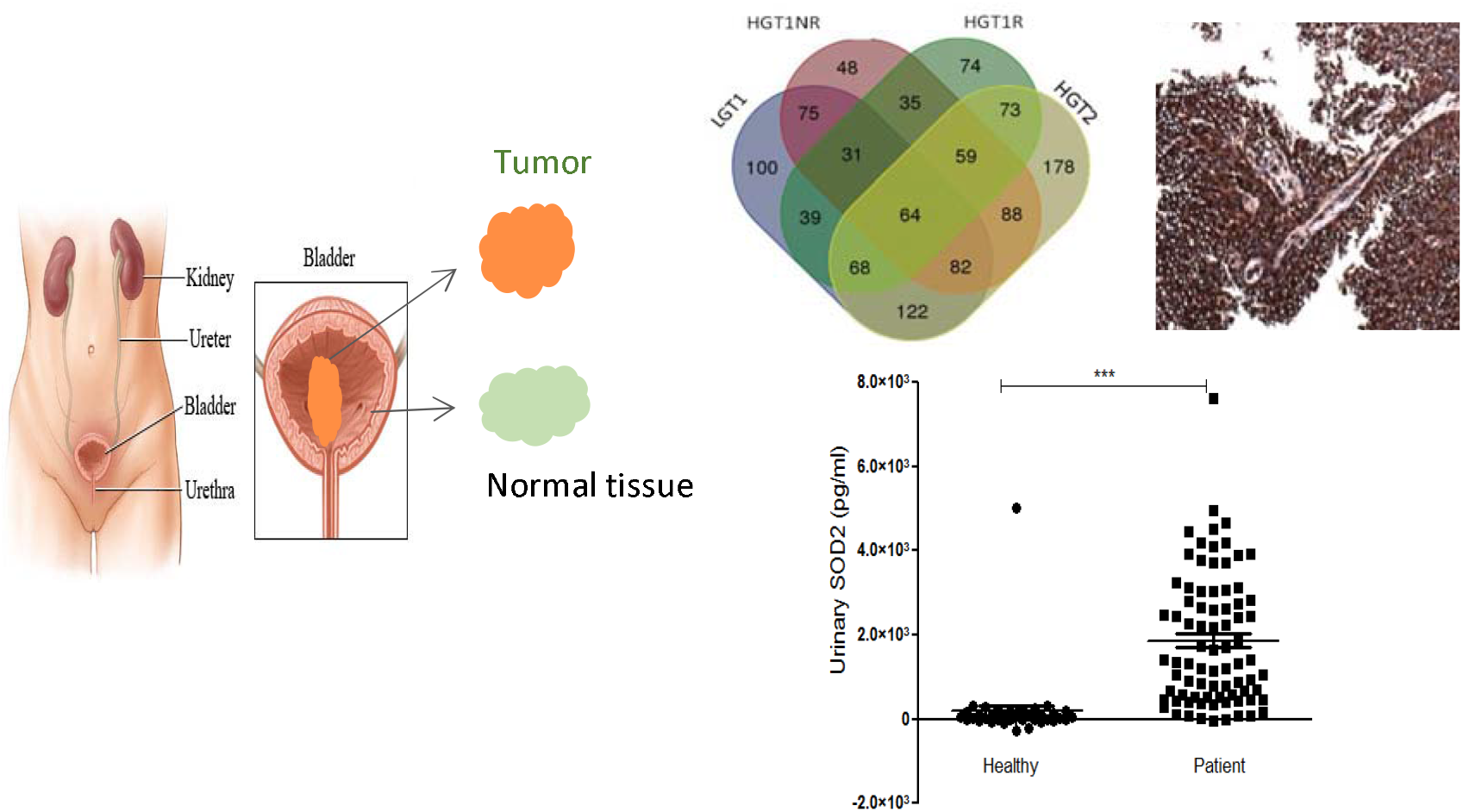

## References

1. https://gco.iarc.fr/today/data/factsheets/cancers/30-Bladder-fact-sheet.pdf (accessed on 11/12/2021)

2. Leal, Jose, et al. “Economic burden of bladder cancer across the European Union.” European urology 69.3 (2016): 438–447.

3. Mariotto, Angela B., et al. “Projections of the cost of cancer care in the United States: 2010–2020.” Journal of the National Cancer Institute 103.2 (2011): 117–128.

4. Kamat, Ashish M., et al. “Bladder cancer.” The Lancet 388.10061 (2016): 2796–2810.

5. Freedman, Neal D., et al. “Association between smoking and risk of bladder cancer among men and women.” Jama 306.7 (2011): 737–745.

6. Pesch, Beate, et al. “Screening for bladder cancer with urinary tumor markers in chemical workers with exposure to aromatic amines.” International archives of occupational and environmental health 87.7 (2014): 715–724.

7. Lucca, Ilaria, et al. “Gender differences in incidence and outcomes of urothelial and kidney cancer.” Nature Reviews Urology 12.10 (2015): 585–592.

8. Chou, Roger, et al. “Urinary biomarkers for diagnosis of bladder cancer: a systematic review and meta-analysis.” Annals of internal medicine 163.12 (2015): 922–931.

9. Lotan, Yair, and Claus G. Roehrborn. “Sensitivity and specificity of commonly available bladder tumor markers versus cytology: results of a comprehensive literature review and meta-analyses.” Urology 61.1 (2003): 109–118.

10. Chen, Guoan, et al. “Protein profiles associated with survival in lung adenocarcinoma.” Proceedings of the National Academy of Sciences 100.23 (2003): 13537–13542.

11. Luque□García, Jose Luis, et al. “Differential protein expression on the cell surface of colorectal cancer cells associated to tumor metastasis.” Proteomics 10.5 (2010): 940–952.

12. Grønborg, Mads, et al. “Biomarker Discovery from Pancreatic Cancer Secretome Using a Differential Proteomic Approach* S.” Molecular & Cellular Proteomics 5.1 (2006): 157–171.

13. Narod, Steven A. “BRCA mutations in the management of breast cancer: the state of the art.” Nature Reviews Clinical Oncology 7.12 (2010): 702–707.

14. Aran D, Camarda R, Odegaard J, Paik H, Oskotsky B, Krings G, Goga A, Sirota M, Butte AJ. “Comprehensive analysis of normal adjacent to tumor transcriptomes.” Nat Commun. 2017 8.1 (2017):1077.

15. Holley, Aaron K., et al. “Manganese superoxide dismutase: beyond life and death.” Amino acids 42.1 (2012): 139–158.

16. Pias, Erin K., et al. “Differential effects of superoxide dismutase isoform expression on hydroperoxide-induced apoptosis in PC-12 cells.” Journal of Biological Chemistry 278.15 (2003): 13294–13301.

17. Mayo, Juan C., Rosa M. Sainz, and Isabel Quiros-Gonzalez. “MnSOD/SOD2 in cancer: The story of a double agent.” Reactive Oxygen Species 5.14 (2018): 86–106.

18. Johnson, Felicity, and Cecilia Giulivi. “Superoxide dismutases and their impact upon human health.” Molecular aspects of medicine 26.4-5 (2005): 340–352.

19. Kinnula, Vuokko L., and James D. Crapo. “Superoxide dismutases in malignant cells and human tumors.” Free Radical Biology and Medicine 36.6 (2004): 718–744.

20. Becuwe, Philippe, et al. “Manganese superoxide dismutase in breast cancer: from molecular mechanisms of gene regulation to biological and clinical significance.” Free Radical Biology and Medicine 77 (2014): 139–151.

21. Termini, Lara, et al. “SOD2 immunoexpression predicts lymph node metastasis in penile cancer.” BMC clinical pathology 15.1 (2015): 1–8.

22. Connor, Kip M., et al. “Manganese superoxide dismutase enhances the invasive and migratory activity of tumor cells.” Cancer research 67.21 (2007): 10260–10267.

23. Hurt, E. M., et al. “Molecular consequences of SOD2 expression in epigenetically silenced pancreatic carcinoma cell lines.” British journal of cancer 97.8 (2007): 1116–1123.

24. Liu, Jingru, et al. “Redox regulation of pancreatic cancer cell growth: role of glutathione peroxidase in the suppression of the malignant phenotype.” Human gene therapy 15.3 (2004): 239–250.

25. Talarico, Maria Cecília Ramiro, et al. “High Expression of SOD2 Protein Is a Strong Prognostic Factor for Stage IIIB Squamous Cell Cervical Carcinoma.” Antioxidants 10.5 (2021): 724.

26. Jin, Honglei, et al. “Divergent behaviors and underlying mechanisms of cell migration and invasion in non-metastatic T24 and its metastatic derivative T24T bladder cancer cell lines.” Oncotarget 6.1 (2015): 522.

27. Hempel, Nadine, et al. “Altered redox status accompanies progression to metastatic human bladder cancer.” Free Radical Biology and Medicine 46.1 (2009): 42–50.

28. Goerlitz, David, et al. “Genetic polymorphisms in NQO1 and SOD2: interactions with smoking, schistosoma infection, and bladder cancer risk in Egypt.” Urologic Oncology: Seminars and Original Investigations. Vol. 32. No. 1. Elsevier, 2014.

29. Nikic, Predrag, et al. “Association between GPX1 and SOD2 genetic polymorphisms and overall survival in patients with metastatic urothelial bladder cancer: a single-center study in Serbia.” J. BUON 23 (2018): 1130–1135.

