## Supplementary material for "Proteomics analysis in urinary bladder cancer patients identifies urinary SOD2 as a predictive marker of recurrence": Figure S1 and Figure S2 and Table S1 and Table S2 and Table S3

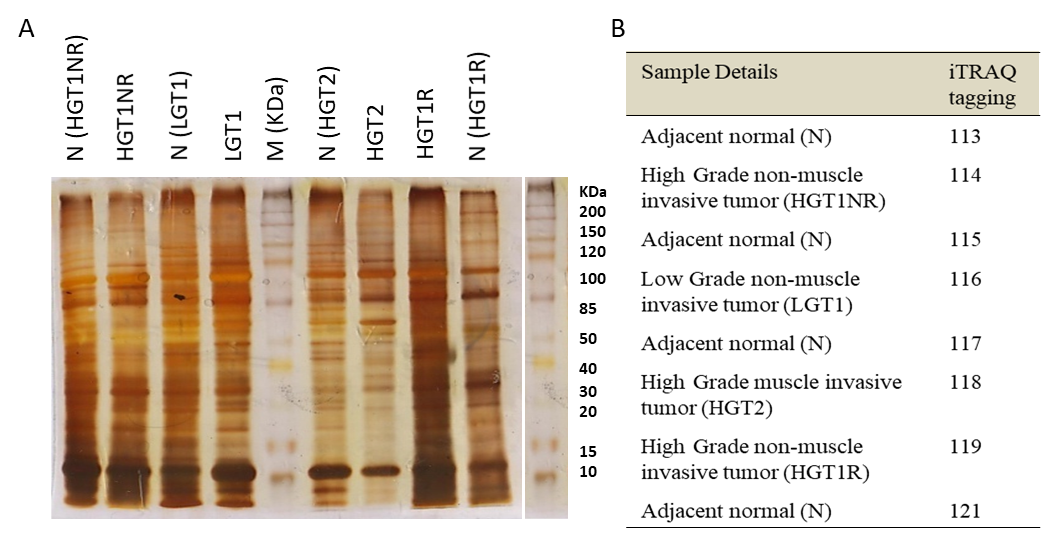

**Figure S1. Selection and iTRAQ tagging of sample in discovery phase.** SDS-PAGE (12.5%) image stained with silver staining of 12 cases included tumor and paired adjacent normal mucosa having 4 µg sample per well showed protein distribution of A) well number 1= 014 N(HGT1NR); 2= 014 HGT1NR; 3=017 N(LGT1); 4= 017 LGT1; 5= protein marker; 6= 020 N(HGT2); 7=020 HGT2; 8= 013 HGT1R; 9= 013 N(HGT1R) used in 1st set and 2nd set of iTRAQ experiment. B) samples used for iTRAQ labeling in discovery phase are shown in figure.

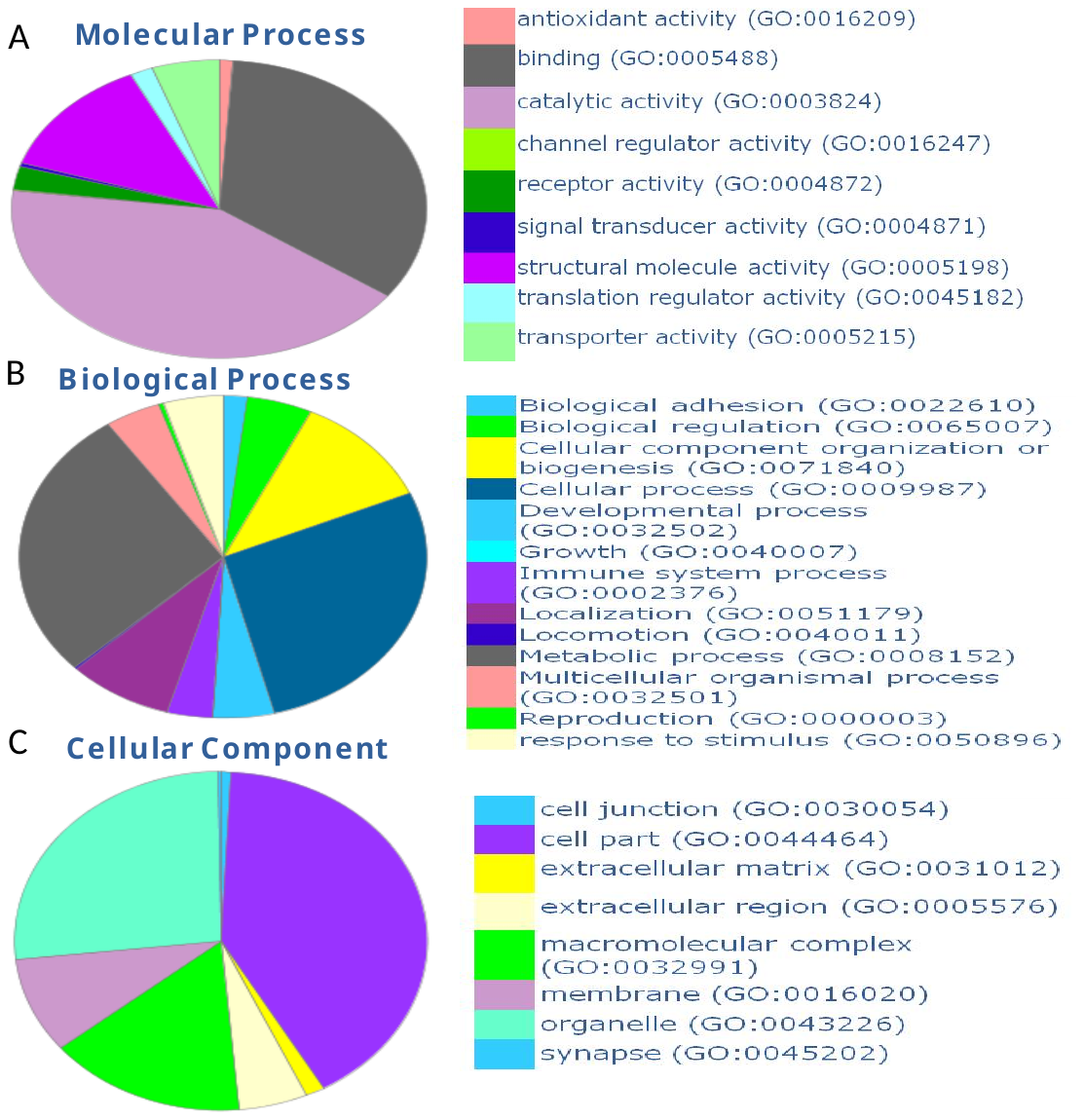

**Figure S2: Determination of Gene ontology (GO) terms of deregulated proteins of subgroups of urotheilal bladder cancer using PANTHER 9.0.** Pie chart is showing 5.5 A) Cellular component (40.9% cell part), B) Biological Process (27.5% cellular and 27.5% metabolic process), C) molecular Process (42% catalytic activity).

**Table S1: Demographic details of independent study cohort of each phase of experiments**

| Demographic details and classification | Total  Study population  (n=212) | Discovery phase | | Verification phases | | | | Validation phase | |
| --- | --- | --- | --- | --- | --- | --- | --- | --- | --- |
|  |  | (iTRAQ)  Paired tissue | | (IHC)  FFPE tissue | | (WB)  Urine | | (ELISA)  Urine | |
|  |  | Tumor tissue  (n=4) | Normal mucosa  (n=4) | Patients  (n=15) | Normal mucosa  (n=3) | Patients  (n=26) | Healthy  (n=10) | Patients  (n=100) | Healthy  (n=50) |
| Median Age  (1^st^ to 3^rd^ IQR) | 58  (47-63) | 61  (56-62) | 61  (56-62) | 58  (53-68) | 54  (51-60) | 58  (52-64) | 53  (48-60) | 58  (49-56) | 55  (45-61) |
| Gender: Female/Male | 43/169 | -/4 | -/4 | 5/10 | -/3 | 4/22 | 1/9 | 21/79 | 12/38 |
| Grade: LG/HG | 6/13 | 1/3 | -- | 5/10 | -- | -- | -- | -- | -- |
| Stage: PT1/PT2 | 13/6 | 3/1 | -- | 10/5 | -- | -- | -- | -- | -- |
| Recurrence | 51 | 1 (25) | -- | -- | -- | 9(35%) | -- | 41(27 %) | -- |

*LG-low grade; HG-High grade; PT1-Non-muscle invasive; PT2-Muscle invasive; iTRAQ-Isobaric tags for relative and absolute quantitation; IHC-Immunohistochemistry; WB- western blot; ELISA-Enzyme-linked immunosorbent assay*

**Table S2: Fold change of common up and downregulated proteins in subgroups of bladder cancer.**

| **Name** | **Protein code** | **LGT1** | **HGT1NR** | **HGT1R** | **HGT2** |
| --- | --- | --- | --- | --- | --- |
| **Up-regulated proteins** | | | | | |
| Sterol-4-alpha-carboxylate 3-dehydrogenase | NSDHL | 2.83 | 1.65 | 1.18 | 1.57 |
| FLJ00385 protein | Q8NF17 | 2.70 | 2.10 | 1.22 | 1.87 |
| Ig kappa chain V-IV | KV402 | 2.64 | 1.17 | 1.18 | 1.89 |
| Caldesmon | CALD1 | 2.51 | 2.64 | 1.05 | 3.87 |
| Plasma protease C1 inhibitor | H9KV48 | 2.27 | 4.05 | 1.81 | 4.62 |
| Collagen type VI, alpha 3 | D9ZGF2 | 1.75 | 3.61 | 1.85 | 4.12 |
| Heat shock protein beta-6 | K7EP04 | 1.66 | 5.14 | 1.58 | 3.52 |
| Superoxide dismutase | Q7Z7M4 | 1.36 | 1.73 | 1.34 | 2.38 |
| Integrin-linked protein kinase | ILK | 1.25 | 1.66 | 3.32 | 3.49 |
| **Down-regulated proteins** | | | | | |
| Calnexin | B4DGP8 | -1.09 | -1.51 | -1.24 | -3.77 |
| Peptidyl-prolylcis-trans isomerase | PPIB | -1.17 | -1.44 | -6.50 | -6.34 |
| 40S ribosomal protein S14 | RS14 | -1.20 | -2.82 | -2.62 | -2.22 |
| Cytochrome b-c1 complex | QCR1 | -1.22 | -1.86 | -1.61 | -1.29 |
| Glycine cleavage system H-protein | Q6QN92 | -1.42 | -1.06 | -3.18 | -3.55 |
| ATP synthase-coupling factor 6 | Q6IB54 | -1.45 | -1.46 | -2.30 | -1.79 |
| Lysosome-associated membrane glycoprotein 1 | LAMP1 | -1.57 | -1.74 | -6.50 | -6.46 |
| Putative uncharacterized protein DKFZp686P17171 | Q63HR1 | -1.66 | -4.62 | -4.45 | -2.88 |
| ATP synthase subunit beta | ATPB | -1.66 | -1.26 | -1.38 | -1.95 |
| Lamin-B1 | LMNB1 | -1.69 | -1.29 | -1.24 | -1.34 |
| Cation-dependent mannose-6-phosphate receptor | Q53GY9 | -1.94 | -2.21 | -3.23 | -4.13 |
| NADH dehydrogenase ubiquinone 1 alpha 4 | NDUA4 | -2.27 | -1.37 | -1.55 | -2.26 |
| Protein S100-P | S100P | -2.39 | -1.37 | -3.97 | -1.14 |
| Transformation-related protein 14 | Q597H1 | -2.54 | -3.28 | -1.17 | -1.05 |
| Lamin A/C transcript variant 1 | Q5I6Y6 | -2.78 | -1.09 | -1.24 | -1.26 |
| Chaperonin 10-related protein | Q9UNM1 | -3.14 | -2.83 | -1.51 | -2.98 |
| cDNA FLJ51907, highly similar to Stress-70 protein | B7Z4V2 | -3.23 | -1.01 | -1.49 | -3.20 |
| cDNA, FLJ94640, highly similar to Homo sapiens keratin 18 | B2RA03 | -3.26 | -1.06 | -2.05 | -1.49 |

**Table S3: Ingenuity network summarizes top diseases and function associated with significantly up/down regulated proteins.**

|  | Molecules in Network | Score | Focus Molecules | Top Diseases and Function |
| --- | --- | --- | --- | --- |
| 1 | 26s Proteasome, AIFM1, AKT, APOA1, CALD1, CANX, ERK1/2, estrogen receptor, FN1, HSPA9, HSPB1, IgG, IGHG1, ILK, Jnk, KRT18, LAMP1, LDL, LMNA, MCAM, MUT, MYLK, NDRG1, NFκB(complex), P38 MAPK, p85 (pik3r), PPIF, PPP1R12A, SAR1A, SERPINA1, SMAD4,Smad2/3**, SOD2**, SRSF3, UBE2N | 51 | 24 | Cellular Movement, Hematological system Immune cell Trafficking |
| 2 | ANGPTL4, ANXA3, ATP5B, ATP5J, CASP4, CHK4, COL12A1, COL6A3, CST5, CXCL8, EPAS1, FBN1, FHL1, Fibrinogen, FSH, GLS, GRB2, HIF1A, **HSPB6**, HSPE1, Lh, LMNB1, LONP1,MAP1LC3B, NRG1, PDLIM3, PRKCZ, PTGIS, S100A11, SERBP1, SF3A3, SFPQ, STK17A, TCR, TGFB1 | 25 | 14 | Nucleic Acid Metabolism, Small Molecule biochemistry, Development Disorder |
| 3 | ACSL1, BRCA1, CDH4, DDB2, F11, FKBP5,G6PD, GF11, H3F3A/H3F3B, HBB, Histone h3, HNF1A, HNF1A, HNF1B, ICAM1, IDH1, IGFBP7, JMJD1C, MTA2, MYC, NDRG1, NR5A2, PGR, RAB27B, REL, RNU7-1, RPS14, S100P, **SERPING1**, SMO, SOD2, SRSF7, TP63, UGT1A7 (includes others), WNT5A, XRCC6 | 18 | 11 | Cell Death and Survival, Cellular Development, Cellular Growth and Proliferation |
| 4 | MiR-124-3P (and other miRNAs w/seed AAGGCAC), SUCLG2 | 2 | 1 | Cellular Function and Maintenance, Lipid Metabolism, Nucleic Acid Metabolism |
| 5 | ERM, SLK | 2 | 1 | Cell Morphology, Cellular Assembly and Organization, Cellular Function and Maintenance |
| 6 | LY6D, TSTA3 | 2 | 1 | Carbohydrate Metabolism, Cell-to-cell Signalling and interaction, Post-Translational Modification |
| 7 | RAC1, SH3BP1 | 2 | 1 | Cell Morphology, Carbohydrate Metabolism Cardiovascular system development and function |
| 8 | KAT5, OGN | 2 | 1 | Cancer, Cell Death and Survival, Cellular Development |
| 9 | MGEA5, NSDHL | 2 | 1 | Developmental Disorder, Hereditary Disorder, Neurological Diseases |
| 10 | miR-145-5p (and other miRNAs w/seed UCCAGUU), NDUFA4 | 2 | 1 | Cancer, Cell Morphology, Cellular Response to therapeutics |
| 11 | RTN4, UQCRC1 | 2 | 1 | Cell Death and Survival, cell morphology, Cellular Assembly and Organization |
